# Identifying COVID-19 cases in primary care settings

**DOI:** 10.1101/2020.08.26.20182204

**Authors:** Yinan Mao, Yi-Roe Tan, Tun Linn Thein, Yi Ann Louis Chai, Alex R Cook, Borame L Dickens, Yii Jen Lew, Fong Seng Lim, Jue Tao Lim, Yinxiaohe Sun, Meena Sundaram, Alexius Soh, Glorijoy Shi En Tan, Franco Pey Gein Wong, Barnaby Young, Kangwei Zeng, Mark Chen, Desmond Ong Luan Seng

## Abstract

Case identification is an ongoing issue for the COVID-19 epidemic, in particular for outpatient care where physicians must decide which patients to prioritise for further testing. This paper reports tools to classify patients based on symptom profiles based on 236 SARS-CoV-2 positive cases and 564 controls, accounting for the time course of illness at point of assessment. Clinical differentiators of cases and controls were used to derive model-based risk scores. Significant symptoms included abdominal pain, cough, diarrhea, fever, headache, muscle ache, runny nose, sore throat, temperature between 37.5°C and 37.9°C, and temperature above 38°C, but their importance varied by day of illness at assessment. With a high percentile threshold for specificity at 0.95, the baseline model had reasonable sensitivity at 0.67. To further evaluate accuracy of model predictions, we firstly used leave-one-out cross-validation, which confirmed high classification accuracy with an area under the receiver operating characteristic curve of 0.92. For the baseline model, sensitivity decreased to 0.56. Secondly, in a separate ongoing prospective study of 237 COVID-19 and 346 primary care patients presenting with symptoms of acute respiratory infection, the baseline model had a sensitivity of 0.57 and specificity of 0.89, and in retrospective notes review of 100 COVID-19 cases diagnosed in primary care, sensitivity was 0.56. A web-app based tool has been developed for easy implementation as an adjunct to laboratory testing to differentiate COVID-19 positive cases among patients presenting in outpatient settings.

## Introduction

The coronavirus disease 2019 (COVID-19) pandemic caused by SARS-CoV-2 poses ongoing challenges for rapid case detection to ensure timely treatment and isolation (1). The disease can progress quickly to acute respiratory distress in severe cases, especially among at-risk groups (2-4). With therapeutic options emerging (5), recognizing cases at milder stages before they deteriorate, usually after the first week of illness (6), can be lifesaving. Moreover, due to the high transmissibility of SARS-CoV-2 in the earlier phases of illness (7), earlier identification can reduce onward transmission (8,9). Ideally, clinicians use reverse transcription polymerase chain reaction (RT-PCR) tests for diagnosis (10) but the caseload and associated testing costs may make this infeasible, particularly in resource-limited regions.

Symptom-based diagnosis is challenging as many of the commonly reported symptoms of COVID-19 are shared by other respiratory viruses, including fever and dry cough (9,11). While several studies have developed algorithms to differentiate COVID-19 from non-COVID-19 patients, almost all were based on static clinical measurements taken at presentation, most commonly to tertiary care facilities (12). However, models that account for symptoms reported at an earlier stage, when patients may first present to outpatient departments, would be advantageous in reducing the delay to treatment and isolation.

To improve case identification, we developed a tool which evaluates patients based on their symptom profile up to 14 days post-onset using a case-control design, with 236 SARS-CoV-2 positive cases evaluated at public hospitals, and 564 controls recruited from a large primary care clinic. We determine the clinical differentiators of cases and controls and apply the algorithm to independent data on cases and controls. We show the importance of incorporating time from symptom onset when deriving model-based risk scores for clinical differentiation of COVID-19 from other patients presenting with symptoms of acute respiratory infection.

## Methods

### Data for model building and independent validation

Anonymized data from controls was prospectively collected in a large public sector primary care clinic in Singapore as part of a quality improvement project from 4 March 2020 to 7 April 2020. We included patients of at least 16 years of age, evaluated by a doctor to be suffering from an acute infectious respiratory infection, and who presented with any of 10 symptoms: self-reported feverishness, cough, runny nose or blocked nose, sore throat, breathlessness, nausea or vomiting, diarrhea or loose stools, headache, muscle ache, abdominal pain. Sequential sampling was adopted wherein the first 5 to 10 eligible patients in each consultation session were recruited. Doctors completed a data collection sheet, which included patients’ age, gender, tympanic temperature, and symptoms since the onset of illness.

Data from COVID-19 cases was obtained from patients admitted to seven public sector hospitals in Singapore, to 9 April 2020. All SARS-CoV-2 was confirmed by RT-PCR testing of respiratory specimens as previously described (13). Demographic data and detailed information on symptoms, signs and laboratory investigations were collected using structured questionnaires with waiver of consent granted by the Ministry of Health, Singapore under the Infectious Diseases Act. We extracted data on the same fields as described above from either the first day of presentation to hospital until they were discharged or until 15 days from illness onset. For both primary care controls and COVID-19 cases, individuals who had temperatures of 37.5°C or above were defined as having a fever. For COVID-19 cases following their admission, any incomplete temperature measurements after their fever end dates were imputed to have a temperature <37.5°C.

Data for independent validation came from two sources. An ongoing prospective study COVID-19 patients admitted to the National Centre for Infectious Diseases quantified sensitivity, while specificity was measured in controls presenting with symptoms of acute respiratory infection recruited at 34 different primary care clinics between 14 March to 16 June 2020. Secondly, to independently assess sensitivity for cases presenting in outpatient care, we extracted data on symptoms used by the model through retrospective chart reviews of patients testing positive for COVID-19 between 17 March to 22 May 2020 at 5 large public sector primary care clinics.

### Modelling for predictive risk using symptom characteristics

We stratified illness days into four intervals for analysis (days 1-2, 3-4, 5-7 and 8 or more) to account for the temporal evolution of symptoms amongst cases and controls; day 1 was the date of symptom onset.

The covariates, with interactions with illness days, were first selected from the candidate list by fitting a logistic regression model to compare cases versus controls. Variables were selected with a least absolute shrinkage and selection operator penalty; covariates with non-zero coefficients were included in subsequent analysis. The analysis used the R package glmnet (14). After variable selection, a second round of unpenalized logistic regression (15) was carried out. Covariates included the interaction terms with illness days for symptoms and temperature in up to 3 categories (<37.5°C; 37.5–37.9°C; ≥38.0°C).

As incrementally more COVID-19 cases present to care over the course of their illness, there are more observations in later dates post illness onset. Contrariwise, the proportion of cases that can potentially present at primary care should decrease as an increasing proportion get diagnosed or recover. As the true distribution of cases presenting to primary care on different days of illness was not available, we assumed that each illness day would result in a linear decrease in the number of cases that can be diagnosed. This was implemented by assigning weights to cases:

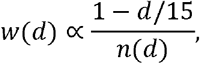

where *d* = 1,2,…,14 represents illness day and n is the count of records in each respective day. Cases remaining undiagnosed on or beyond 15 days after onset of symptoms are considered to have recovered (16).

### Performance evaluation

The performance of the model was evaluated by the area under the curve (AUC), or the area under the receiver-operating-characteristic (ROC) curve using leave-one-out-cross-validation (LOOCV) to alleviate issues with overfitting.

### Optimising classification of COVID-19 cases and validation on independent datasets

We proposed two strategies to determine the cutoff for differentiating COVID-19 positive or negative patients which prioritised high specificity (e.g. > 0.95) with satisfactory sensitivity (e.g. ~0.7). High specificity was prioritised to limit wrongly classified controls to a level within the capacity of resources available for testing patients. The first strategy obtains a single cutoff with a minimum threshold for overall specificity. The minimum specificity above the threshold corresponding to the best sensitivity on the ROC using full dataset is chosen (17). The second strategy creates multiple cutoffs across illness days with a minimum threshold of overall specificity. A cutoff is chosen for each illness day group, which gives an overall specificity that meets this minimum threshold. For illness day 1-2, 3-4, 5-7 and 8 or more, an optimal combination of cut-offs was determined using a stochastic search algorithm. Classification results are presented using observations from all cases, and separately for 223 observations from 26 cases severe enough to need admission to an intensive care ward, using the full dataset and on LOOCV. To assess potential degradation of performance when applying model to other outpatient settings, we also tested all classification strategies on the independent validation datasets.

## Results

The model building dataset included single-day observations from 564 patients assessed at primary care clinics, and 236 COVID-19 patients admitted to hospital who contributed a median of 6 and a total of 1466 observations on symptoms and body temperature. The independent validation datasets included 237 COVID-19 patients and 346 controls from the prospective study, and 100 COVID-19 patients from retrospective chart reviews. Patient profiles are in Table 1.

**Table 1.**
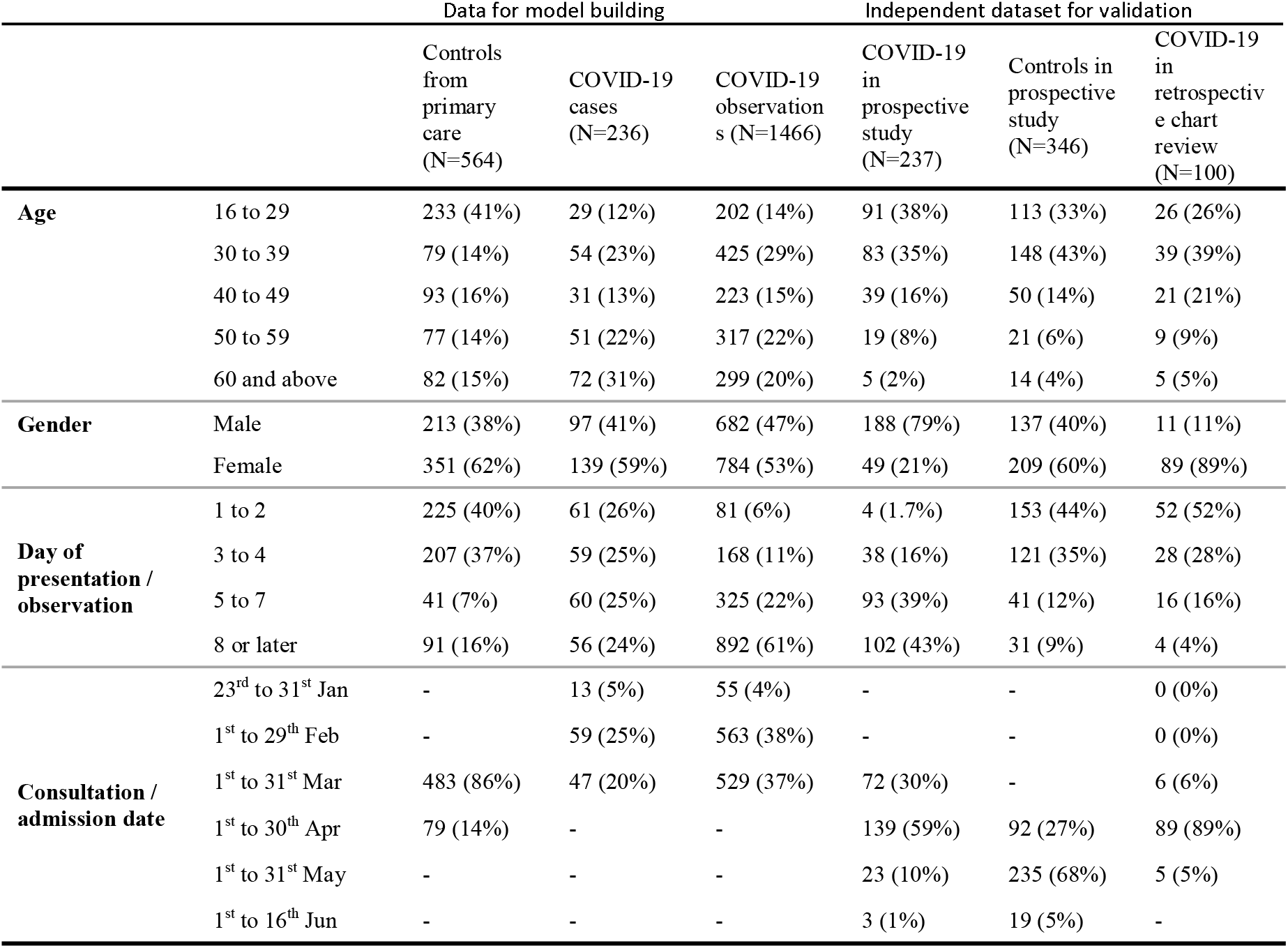
Demographic and clinical profile of case and control patients in 2020 used for model building and for model validation

### Clinical Characteristics of Patients used in model building

SARS-CoV-2 positive cases differed from controls in the proportions presenting with different symptoms over time (Figure 1). Overall, while only a slightly larger proportion of controls ever had cough, a runny nose, sore throat and headache were substantially more common than in SARS-CoV-2 positive patients (Figure 1). A larger proportion of COVID-19 positive patients ever had fever, diarrhea, nausea and/or vomiting, and higher tympanic temperatures. Notably, among COVID-19 patients, the proportions who ever had shortness of breath (SOB), diarrhea and nausea and/or vomiting increased over the course of the illness, while those with temperatures ≥37.5°C and ≥38°C became fewer.

**Figure 1:**
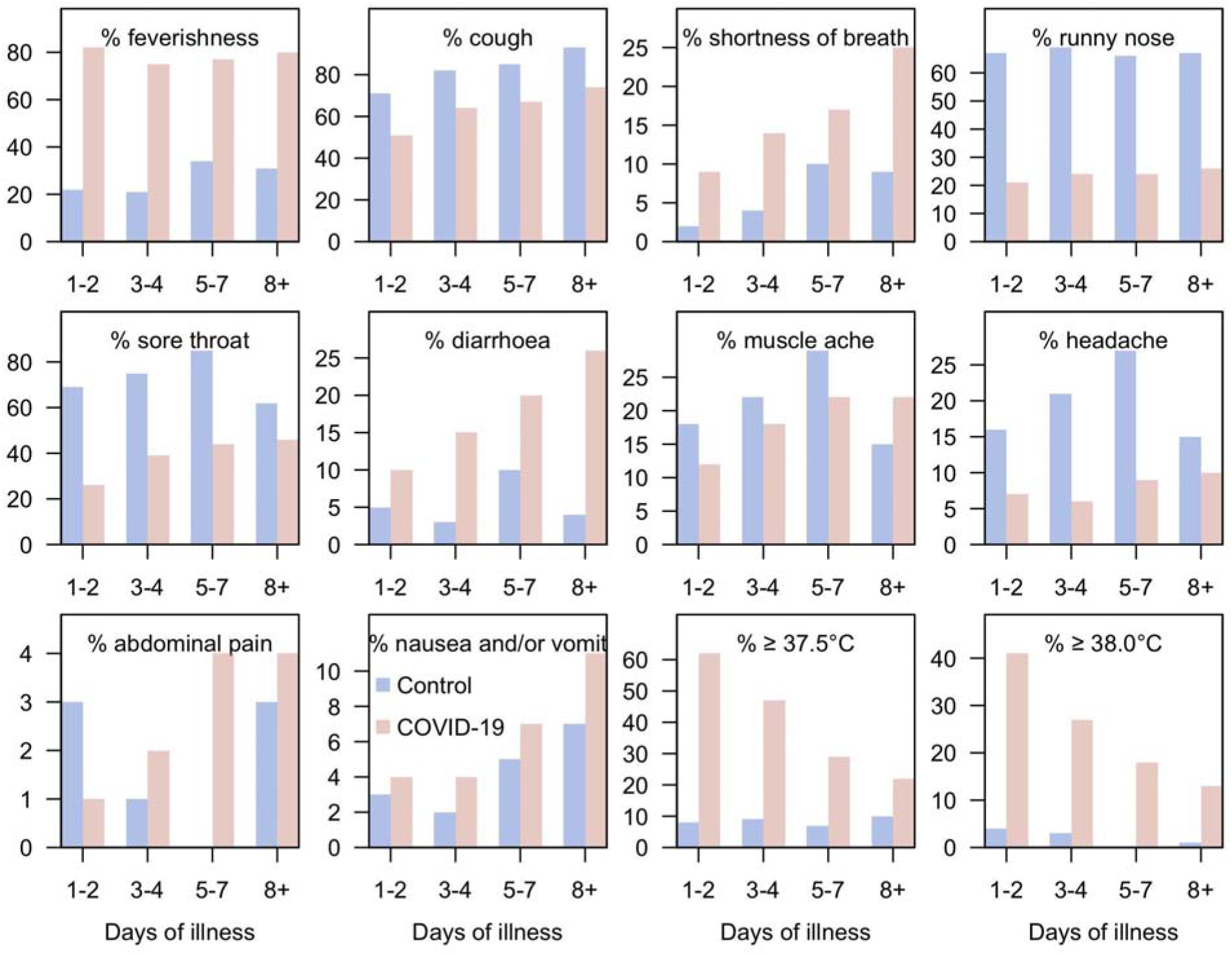
Proportion of cases with described symptoms across days of illness.

### Modelling symptom cutoffs

Figure 2 shows adjusted odds ratios (ORs) for symptoms and illness days (1-2, 3-4, 5-7, 8+). Temperature readings between 37.5°C and 37.9°C significantly increased the OR by 10.89 (95% CI: 3.91–30.35) on illness day 1–2, as well as on illness day 3–4 (OR: 2.55, 95% CI: 1.19–5.48) but not thereafter, and likewise for temperature readings ≥38.0°C (day 1–2 OR: 9.04, 95% CI: 3.78–21.60, day 3–4 OR: 3.02, 95% CI: 1.21–7.53). Ever feeling feverish was consistently associated with SARS–CoV–2 (on day 1–2: OR 7.47, 95% CI: 4.06–13.81, on day 3–4: 8.85, 95% CI: 5.10–15.33, on day 5–7: 10.38, 95% CI: 5.23–20.58, on day 8 or more: 7.12, 95% CI: 4.05–12.55). Diarrhea had a significantly increased OR on all illness days except day 1–2 (on day 3–4: 3.38, 95% CI: 1.24–9.22, on day 5–7: 5.03, 95% CI: 1.59–15.89, on day 8 or more: 10.58, 95% CI: 3.43–32.67).

Runny nose was consistently associated with decreased OR (on day 1–2: 0.13, 95% CI: 0.07–0.22, on day 3–4: 0.21, 95% CI: 0.13–0.35, on day 5–7: 0.28, 95% CI: 0.14–0.53, on day 8 or more: 0.12, 95% CI: 0.07–0.21). Abdominal pain in the early stage of illness on day 1–2 was associated with a lower OR (0.07, 95% CI: 0.01–0.44), as was muscle ache on day 1–2 (OR: 0.21, 95% CI: 0.08–0.53), but not thereafter. Having a headache associated with lower OR when presenting on day 3–4 (OR: 0.23, 95% CI: 0.11–0.49), and on day 5–7 (OR: 0.40, 95% CI: 0.17–0.96). Having a sore throat was also negatively associated with COVID–19 on day 1–2 (OR: 0.14, 95% CI: 0.08–0.25), day 3–4 (OR: 0.28, 95% CI: 0.18–0.44).

**Figure 2:**
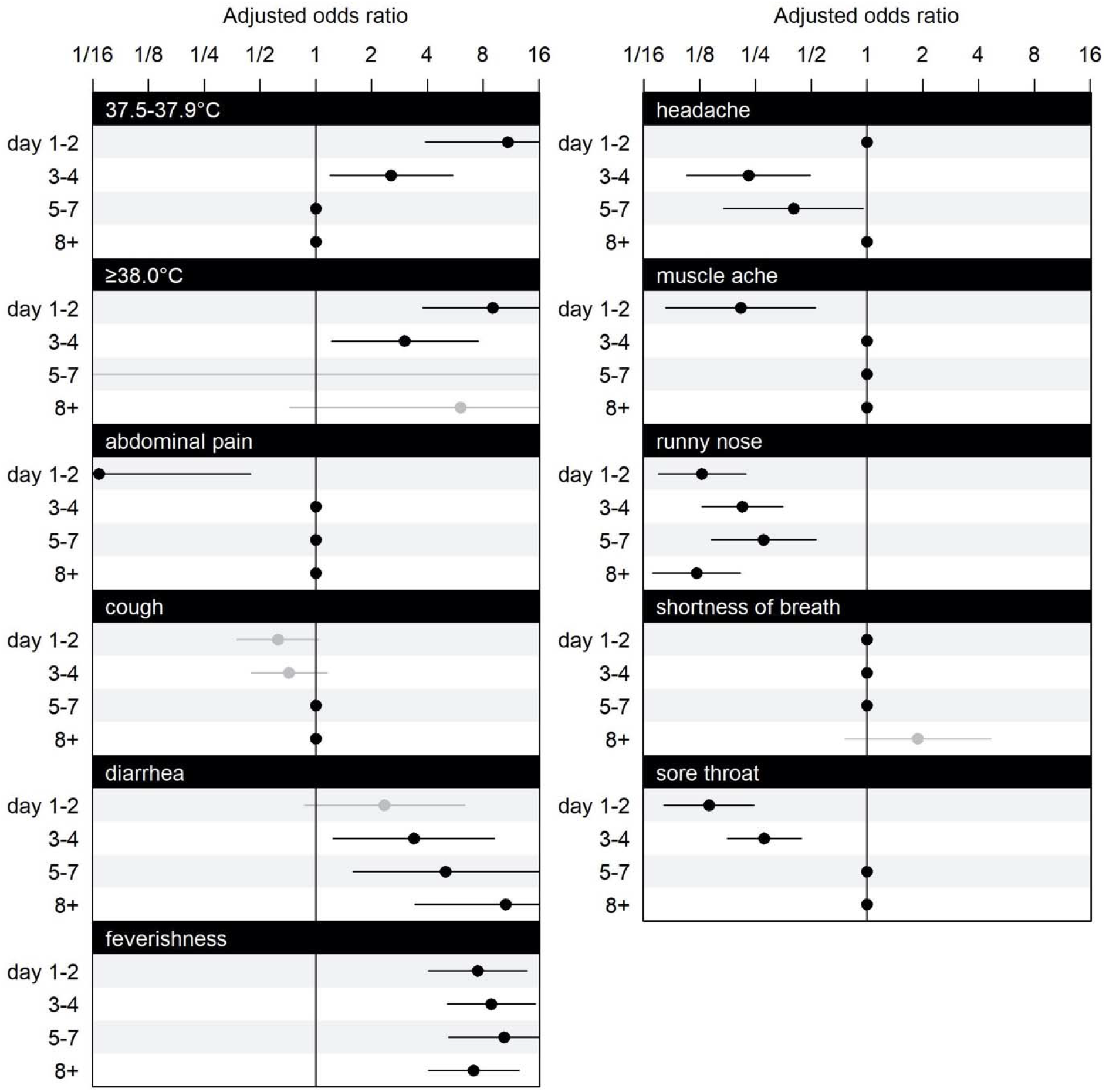
Estimated coefficient means and confidence intervals in the GLM. The dots indicate the mean increase in odds ratio for the respective linear effect, and line segments are shown as confidence intervals. Confidence intervals coloured in black indicates significant effects, and grey indicates non-significant effects. Variables with only mean at 1 and without confidence interval were pre-excluded by GLM Lasso and are not included in modelling. Having nausea or vomiting, is omitted from the figure because all of its interaction effects with illness days are excluded by GLM lasso. The scale of parameter effect on the odds ratio is exponentially spaced for visualization.

### Predictive Performance

The model had an AUC of 0.89 on LOOCV (Figure 3). With predicted scores above 0.95, 97.9% of the sample are COVID-19 cases (Figure 4a). For predicted scores below 0.2, the proportion which were COVID-19 positive cases was 3.9%. Reasonable separation of the predicted scores was observed between cases and control (Figure 4b). The calculated sample sensitivity, specificity, positive predicted value (PPV) and negative predicted value (NPV) across a grid of cutoffs between 0 and 1 are in Figure 4c.

**Figure 3:**
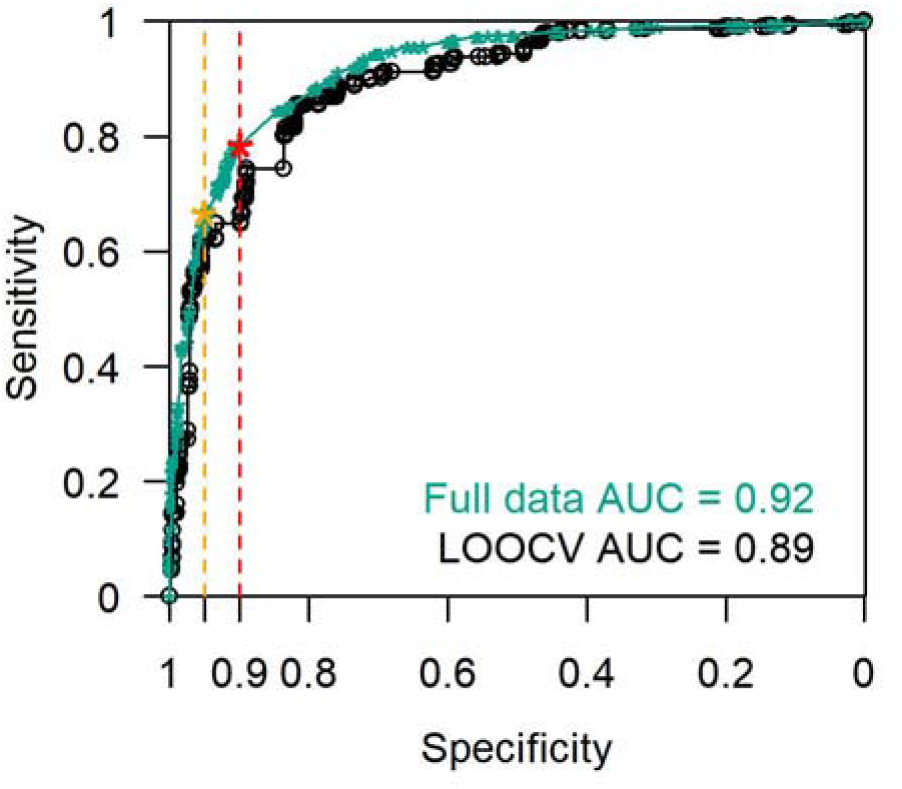
ROC curve with LOOCV. AUC = 0.89. Using full data, AUC = 0.92. With a minimum specification threshold at 0.95 and 0.9, the cutoff points are found at 0.92 and 0.74 respectively as indicated by the orange and red stars on the curve.

**Figure 4:**
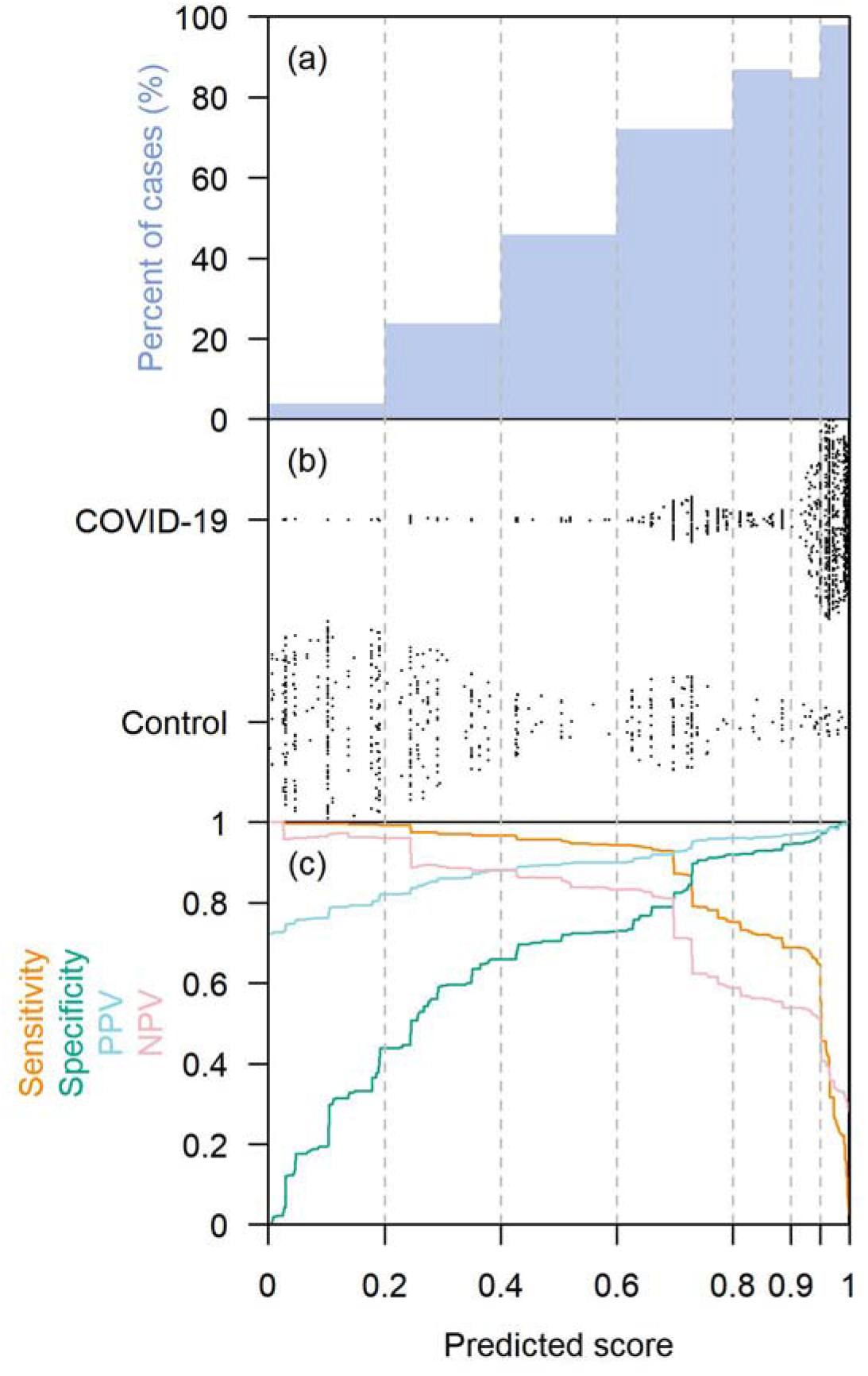
Comparisons of predicted risk to observations, (a) shows bars with the height indicating the percentage of cases in respective intervals of predicted risk. (b) plots the predicted risk grouped by case or control. (c) traces the calculated in-sample sensitivity, specificity, PPV and NPV for a grid of predicted risks as cutoffs spaced at 0.001 from 0 to 1.

### Classification cutoffs

Table 2 gives results from implementing various cutoffs with target specificities of 90 to 95% using the two proposed strategies (see methods). The model performed better for severe cases with overall detection rate at 88% for strategy 1 (with a single threshold for all illness days), and 83% for strategy 2 for a target specificity ≥95%. With a relaxed 90% minimum specificity threshold, the detection rates for severe cases were 98% for strategy 1, and 96% for strategy 2.

**Table 2:** The number of cases detected and its percentage by illness days with cutoff schemes by strategy 1 and 2 at an overall cutoff of 0.95 minimum specificity threshold, and 0.9 minimum specificity threshold, and performance on the independent validation datasets. Strategy 1 chooses a single cutoff point on the LOOCV ROC curve that meets minimum specificity threshold, and strategy 2 searches for an optimal combination of cut-offs for each illness day group that gives an overall specificity that meets this minimum threshold, using a stochastic search algorithm.

|  |  | With 95% specificity |  | With 90% specificity |  |
| --- | --- | --- | --- | --- | --- |
|  | Day | Strategy 1 | Strategy 2 | Strategy 1 | Strategy 2 |
| **Sensitivity using full dataset (n = 2030 cases)** | 2 | 55 (4%) | 62 (4%) | 63 (4%) | 58 (4%) |
|  | 3 | 93 (6%) | 89 (6%) | 122 (8%) | 111 (8%) |
|  | 5 | 221 (15%) | 221 (15%) | 272 (19%) | 308 (21%) |
|  | 8 | 638 (44%) | 362 (25%) | 703 (48%) | 703 (48%) |
|  | Overall | 1007 (69%) | 734 (50%) | 1160 (79%) | 1181 (80%) |
| **Sensitivity for severe cases (n = 223 cases)** | 2 | 10 (4%) | 11 (5%) | 11 (5%) | 10 (4%) |
|  | 3 | 17 (8%) | 17 (8%) | 22 (10%) | 18 (8%) |
|  | 5 | 40 (18%) | 40 (18%) | 43 (19%) | 43 (19%) |
|  | 8 | 130 (58%) | 118 (53%) | 143 (64%) | 143 (64%) |
|  | Overall | 197 (88%) | 186 (83%) | 219 (98%) | 214 (96%) |
| **Sensitivity** | Overall | 67% | 60% | 79% | 79% |
| **Specificity** | Overall | 95% | 96% | 90% | 90% |
|  | F-measure | 0.80 | 0.75 | 0.88 | 0.88 |
| **Validation using LOOCV** | Sensitivity | 56% | 61% | 80% | 74% |
|  | Specificity | 96% | 96% | 83% | 87% |
|  | F-measure | 0.72 | 0.76 | 0.89 | 0.85 |
| **Validation through prospective study (n = 237 cases and 346 controls)** | Sensitivity | 57% | 55% | 78% | 79% |
|  | Specificity | 89% | 88% | 83% | 81% |
|  | F-measure | 0.70 | 0.68 | 0.80 | 0.80 |
| **Validation on retrospective notes review (n = 100 cases)** | Sensitivity | 56% | 62% | 71% | 68% |

Validation using the prospective study caused little change in sensitivity for strategies 1 and 2 at target specificities >90%, but a decrease in sensitivity of 10% and 5% respectively for a target specificity of >95%.

There was also a decrease in observed specificity of between 6 to 9% across all 4 combinations. Using the data from retrospective chart review, we observed 11% drop in sensitivity for both strategy 1 at 95% specificity (from 67% to 56%) and strategy 2 at 90% specificity (from 79% to 68%), with a slight increase of 2% for strategy 2 at 95% specificity and decrease of 8% for strategy 1 at 90% specificity.

## Discussion

Our findings demonstrate the importance of utilizing the symptom profile of COVID-19 across time. Notably, some symptoms were highly differentiated in the proportions observed for cases versus controls, conditional on the time since onset (Figure 1). These differences informed a model that is reasonably discriminatory and specific while retaining good sensitivity. In resource scarce regions, utilizing critical symptom cutoffs as a function of time from symptom onset can facilitate rapid diagnosis where test kits are constrained. Model performance was superior for severe cases in need of more pro-active management. The model’s parsimony facilitates adaptation into a simple tool to risk stratify patients based on reported symptoms and their day of illness onset, as we have done (URL).

We observed COVID-19 symptoms largely in line with previous studies. Cough, breathlessness and fever were present in the clinical presentation of a large proportion(11,12,18), but we must point out how cough has little discriminatory value against other primary care consults and breathlessness is a late symptom associated with more severe illness (15.1% in non-severe cases and 37.6% in severe cases). Our work concurs that feverishness (88.7%) is a dominant symptom, but diarrhea was less common in some other studies (<8.9% in 19–21). Some studies, particularly of hospitalized patients, do suggest the majority would have feverishness (>66.9% in 19–22), and higher proportions with gastrointestinal complaints (~26–37% in 23–25). Differences between studies may be attributable to the inclusion of patients at varying stages of their illness: studies based on hospitalized cases would include more patients at later disease stages, with higher proportions having breathlessness and diarrhea. In our study, diarrhea was not common in early illness but increased in proportion and discriminatory value as the disease progressed. Early presentation of fever and cough is supported in studies (26,27), SOB is presented later at 7 days (26) or 5 days (27), which concur our findings of symptoms through the course of illness (Figure 1). On the other hand, while the importance of feverishness as a symptom has been emphasized, we caution that in a large proportion of COVID-19 cases, the proportion with temperatures ≥37.5°C is only slightly over 60% on D1–2, then drops below 30% from D5 onwards. It decreases in discriminatory value in later illness, and at a stage when a patient may still be infectious.

Compared to existing predictive models for diagnosing COVID-19 patients from symptomatic patients as reviewed in (12), the absence of laboratory and radiographic investigations, and even medical measurements besides body temperature (e.g. blood pressure, oxygen saturation or clinical signs in (3,9,20,27-31), makes our diagnosis tool easier to implement in outpatient practice. Notably, none of the existing diagnostic models in our review of published work account for how illness days modifies the predictive value of different symptoms, though some accounted for the effect of illness day in the variable selection process (e.g. by restricting the analyses to earlier infections (3,20,25,28)). We intentionally omitted demographic and epidemiologic variables as predictors, given the propensity of such associations to change over the course of an epidemic. In spite of this, our model has one of the highest areas under the curve, even on LOOCV, amongst those that do not rely on laboratory investigations. Its performance was still respectable when validated in the prospective study described.

This study has several limitations. Firstly, the distinction between dry and productive cough, and anosmia as a symptom were not captured in this study, particularly because the latter was reported only after our study was started. These have been reported as clinically relevant characteristics for SARS-CoV-2 positive individuals (2,32,33), and their inclusion may have improved the performance of the algorithm further. Secondly, our controls were not tested for COVID-19. However, there was no widespread transmission of COVID-19 at the time of data collection for our controls. For instance, testing of 774 residual sera samples collected in early April 2020, around the time we ceased collecting data controls, identified no seropositive individuals (unpublished data). Thirdly, we recognize that the “controls” against which our COVID-19 patients must be distinguished may differ due to variations in the epidemiology of background illnesses by place and time. This limitation can potentially be overcome by collecting, then repeating the analyses using, updated data from locally relevant “control patients”, collected through the simple data collection format we used.

Our study provides a tool to discern COVID-19 patients from controls using symptoms and day from illness onset with good predictive performance. It could be considered as a framework to complement laboratory testing in order to differentiate COVID-19 from other patients presenting with acute symptoms in outpatient care.

## Data Availability

All data and codes referred to in the manuscript can be shared upon request.

https://sshsphdemos.shinyapps.io/covid-19_calculator_v5/

## Acknowledgement

We would like to acknowledge Prof. Leo Yee-Sin, Dr. Shirin Kalimuddin, Dr. Seow Yen Tan, Dr. Surinder Kaur MS Pada, Dr. Louisa Jin Sun and Dr. Parthasarathy Purnima and the rest of the PROTECT Study group for their contributions.

